# Patient and family engagement interventions in primary care patient safety: a systematic review and meta-analysis

**DOI:** 10.1101/2023.12.29.23300648

**Authors:** Yan Pang, Anna Szücs, Ignacio Ricci Cabello, Jaheeda Gangannagaripalli, Lay Hoon Goh, Foon Leng Leong, Li Fan Zhou, Jose M. Valderas

**Affiliations:** Department of Family Medicine, National University Health System, Singapore; Alice Lee Centre for Nursing Studies, Yong Loo Lin School of Medicine, National University of Singapore; Department of Family Medicine, Yong Loo Lin School of Medicine, National University of Singapore; Balearic Islands Health Research Institute (IdISBa) & CIBER de Epidemiología y Salud Pública (CIBERESP), Spain; NIHR ARC GM /Healthy Ageing research group, School of Health Sciences, The University of Manchester, Manchester, UK; Department of Statistics & Data Science, National University of Singapore, Singapore; Faculty of Behavioural and Movement Sciences, Vrije Universiteit Amsterdam, Netherlands; Centre for Research in Health Systems Performance, National University of Singapore, Singapore, Singapore

**Keywords:** Patient involvement, patient engagement, family involvement, family engagement, patient safety, primary care, family medicine, randomized controlled trial

## Abstract

**Importance:** Engaging patients and their families has been proposed and promoted as a key strategy for improving patient safety of health systems. However, little is known about the use of this approach in the primary care settings.

**Objective:** This systematic review and meta-analysis assessed the effectiveness of interventions promoting patient and family engagement for patient safety in primary care based on randomised controlled trials (RCTs).

**Data Sources:** Five electronic databases (MEDLINE, CINAHL, Embase, Web of Science, and CENTRAL) were searched from inception to February 2023 with key words structured in four blocks (patient and family engagement; patient safety; primary care; randomised controlled trial).

**Study Selection:** Definition of patient safety included adverse events and non-recommended practices. Two independent study team members screened each record, with discrepancies resolved by consensus.

**Data Extraction and Synthesis:** Reporting followed PRISMA standards and included risk of bias and level of certainty assessments. For studies reporting on similar safety outcomes, results were combined into meta-analyses using multi-level random-effects models in case of moderate/substantial heterogeneity (30%≤I²≤75%), and fixed-effect models when heterogeneity was low (I²≤30%).

**Main Outcome(s) and Measure(s):** Expected primary study outcomes were adverse events, non-recommended medical practices, and medical errors. Interventions were considered of interest, if they prompted patients and/or families to take actions, focused on patient education about engagement, or had a significant patient engagement component if they were multifaceted interventions. Interventions were rated based on increasing degrees of patient/family engagement as “Inform about engagement”, “Empower”, and “Partner/Integrate”.

**Results:** Sixteen records were identified, among which eight completed RCTs. No intervention reached the highest engagement level. RCTs primarily targeted medication safety outcomes, with meta-analyses showing no significant effects on adverse drug events (OR=0.73, 95%CI [0.46,1.15]) and medication appropriateness using categorical (OR=0.97, 95%CI [0.73,1.17]) and continuous outcome variables (MD=0.56, 95%CI [-0.61, 1.72]). Overall risk of bias was low and the certainty of evidence ranged from moderate to high for most completed studies.

**Conclusion and Relevance:** Patient and family engagement strategies in primary care show inconclusive results based on extant randomised controlled evidence. They should delve into more comprehensive levels of engagement and address more diverse patient safety outcomes.

**Key points:**

- **Question:** Is there randomised controlled evidence supporting the use of patient and family engagement interventions in primary care patient safety?
- **Findings:** Randomised controlled interventions targeting patient safety through patient and family engagement are scarce in primary care, mostly focus on medication safety, and stay at low to intermediate levels of patient and family engagement. Although their combined effectiveness did not reach significance in meta-analyses, favourable results were reported for several patient safety outcomes.
- **Meaning:** Patient and family engagement interventions for patient safety in primary care show inconclusive results based on the randomised controlled evidence at hand, yet their scarcity and relatively low level of patient/family engagement underscores the need to further test and refine such approaches in all patient safety domains.

## 1 Introduction

Patient safety has gained momentum in the last decades, with patient safety strategies being integrated into the agendas of healthcare organizations worldwide (1). Nevertheless, the development of strategies and interventions to improve patient safety in healthcare delivery has by and large been confined to hospital care (2, 3). As the delivery of care within community settings relies more heavily on patients and their families (4), making them well-placed to identify errors or potential harm risks, strategies targeting patient and family engagement hold particular relevance in primary and community care (5, 6).

Patient and family engagement strategies go beyond raising awareness about care safety and can encompass partnerships between patients/families and healthcare professionals aimed at preventing or mitigating adverse events (1). Even though such strategies were already considered a pillar of patient safety in the landmark report of the Institute of Medicine in 1999, *To Err is Human* (7), only more recently was their importance reemphasized by the World Health Organisation’s Declaration of Astana (8) and the designation of “Engaging patients for patient safety” as the theme for the 2023 World Patient Safety Day (9). The Organisation for Economic Co-operation and Development has recently estimated that effective patient involvement could potentially diminish harm by up to 15% in ambulatory care, leading to significant cost savings for the healthcare system (10). Primary care is an optimal setting to implement such strategies because of the sustained relationship among care providers, patients, and families that is traditionally at its root (11).

Specific interventions, such as face-to-face coaching sessions in older adults (12), family carer support in dementia (13), and the utilization of eHealth tools for reporting adverse drug effects (14), have demonstrated efficacy in engaging patients and families in primary care patient safety. Yet, some authors viewed the implementation of these strategies as challenging (2), with the majority of studies focused on medication safety (14–16). Additionally, numerous patient safety strategies that involve patient/family engagement including patient-provider partnerships (17, 18), patient involvement in decision-making (19), decision coaching (20), patient access to medical records (21), and patient-mediated interventions (22), remain underexplored in primary care. Consequently, it remains uncertain which patient and family engagement interventions are reliably effective in primary care and for which patient safety outcomes (23).

The present systematic review and meta-analysis aims to integrate extant randomised controlled evidence regarding patient and family engagement interventions targeting patient safety outcomes in primary care.

## 2 Methods

We conducted a systematic review following the PRISMA guidelines. The search protocol was preregistered on PROSPERO (registration number: CRD42023397495).

We used Coulter’s definition of patient engagement (24): “*a set of reciprocal tasks between patients, healthcare professionals, and healthcare organizations working together to promote and support active patient and public involvement in health and healthcare and to strengthen their influence on healthcare decisions, at both the individual and the collective level*” (25) and extended it to include patients’ families. Patient safety was defined as “*a health care discipline that aims to prevent and reduce risks, errors and harm that occur to patients during provision of health care*” (26).

We included randomised controlled trials (RCTs) and cluster-randomised trials that recruited participants in primary care settings, such as private practices, family medicine clinics, and community/ambulatory care settings associated with general practice. To be eligible, interventions needed to (i) prompt patients and/or families to take actions in the context of their care; (ii) focus on patient/family education about engagement (e.g., informing about red flags to be signalled to providers); or (iii) have patient/family engagement as a component of a complex intervention, as long as it was reported on separately and involved comparable resources to other components. We only considered safety-related outcomes, such as adverse events leading to increased morbidity/mortality or risk of harm and non-recommended medical practices, such as inappropriate prescriptions. We relied on the authors’ definitions of these terms, given their varying definitions in the literature We excluded non-English language studies, review papers, and conference abstracts, trials of secondary or tertiary healthcare and specialist outpatient care, interventions exclusively involving healthcare providers or policymakers, and outcomes pertaining to quality of care but not explicitly to patient safety.

Five electronic databases were searched for potentially eligible studies, including MEDLINE Ovid, CINAHL EBSCO, Embase Ovid, Web of Science Core Collection, and Cochrane Central Register of Controlled Trials (CENTRAL) in the Cochrane Library. We also performed reference tracking to check for additional eligible records. The search strategy encompassed four blocks: patient and family engagement, patient safety, primary care, and randomised controlled trials (see Supplemental Tables S1 to S5 for the complete search strategy).

Article screening and data extraction were performed by two independent team members for each article, with discrepancies resolved through consensus meetings. Data extraction followed the Cochrane data collection guidelines (27) and risk of bias assessment employed the Cochrane Risk of Bias tool version 2 (28). Certainty of evidence was appraised following the GRADE approach (29).

Records reporting on pilot and full-scale RCTs were grouped by outcomes (continuous versus categorical outcomes) and conceptual similarity (inappropriate prescriptions, side effects, others). For groups containing two or more studies, results were combined in meta-analyses using R’s *metafor* (30) and *meta* (31) packages. In alignment with Cochrane guidelines (32), the longest follow-up time was used. Random-effects models following the DerSimonian and Laird method were built for groups where 30% ≤ I^2^ ≤ 75%, indicating moderate to substantial heterogeneity according to the Cochrane guidelines (33), whereas fixed-effects models following the Mantel-Haenszel method were built for groups with no to negligible heterogeneity (I^2^ ≤ 30%). In the event any group displayed considerable heterogeneity (I^2^ ≥ 75%), it was not combined into a meta-analysis. For categorical outcomes, the analysis employed the natural logarithm of odds ratios and corresponding variance to estimate pooled odds ratios. Meta-analysis of continuous outcomes was reported as mean difference in scores (MD).

Records unsuitable to be combined by meta-analysis were summarised through narrative synthesis. We employed an adapted version of the engagement framework developed by Kim and colleagues (34) to appraise the level of engagement of patients/families in each intervention (Table 1).

**Table 1:**
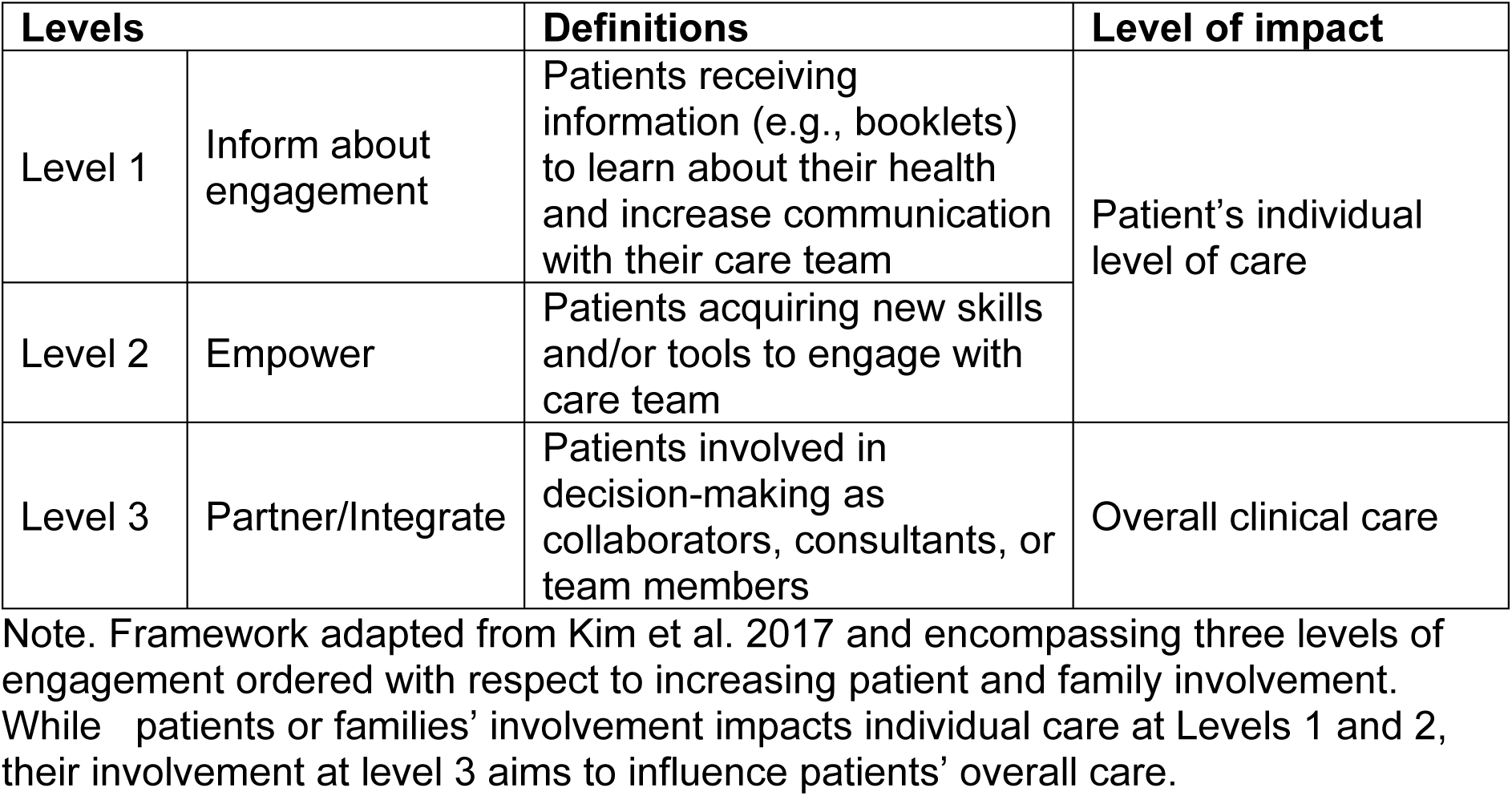
Levels of patient and family engagement.

## 3 Results

### 3.1 Study selection and general characteristics

The systematic search yielded a total of 4,773 records, of which 3,137 remained after deduplication and 173 after full text retrieval (Figure 1). A final set of 16 records were included (Table 2), of which eight were completed RCTs. Raw outcome data was not reported for one study (35), which had to be excluded from the meta-analysis part after two unsuccessful attempts to contact the authors by email.

**Figure 1.**
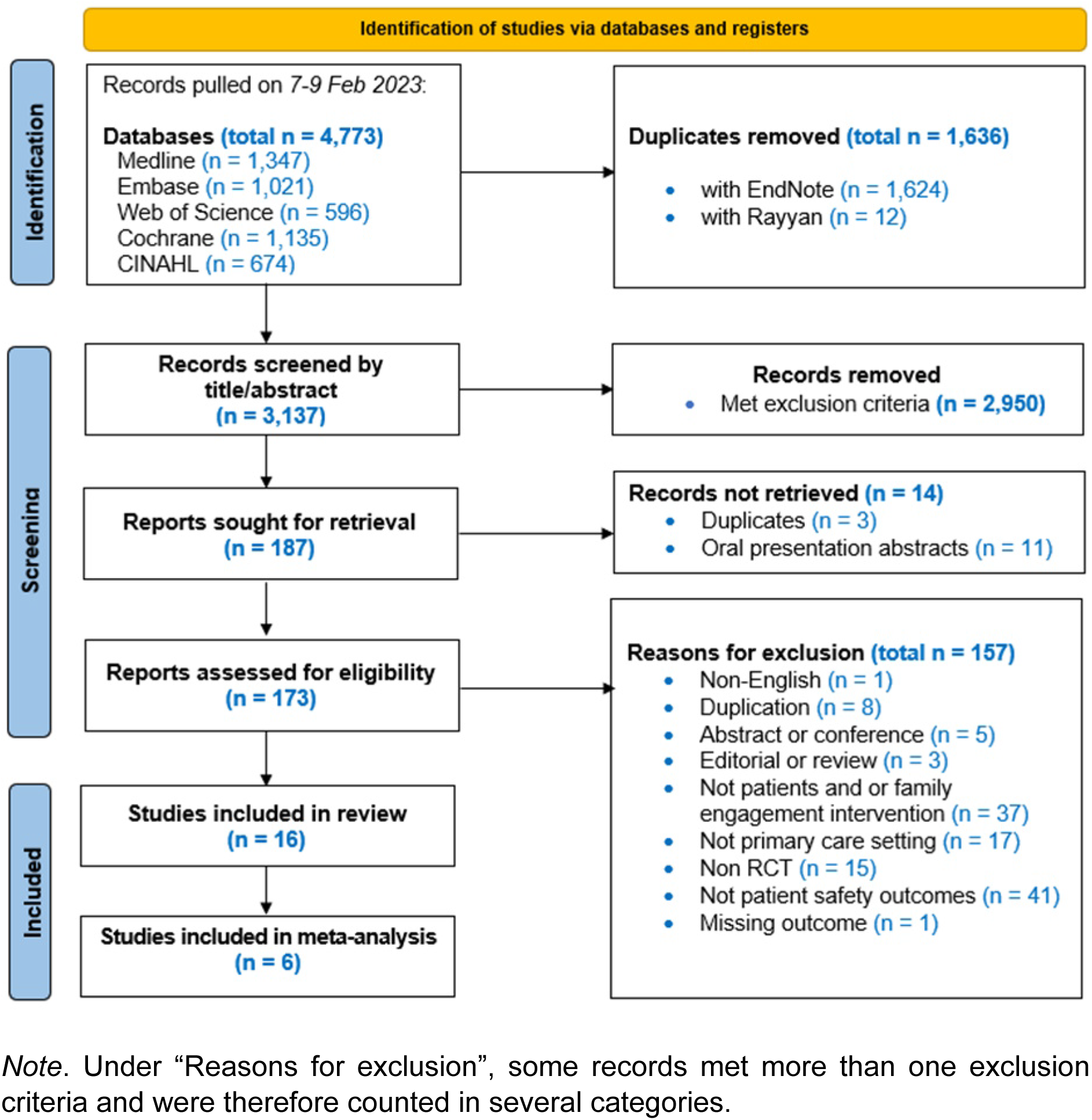
PRISMA Chart summarising the screening process.

**Table 2:**
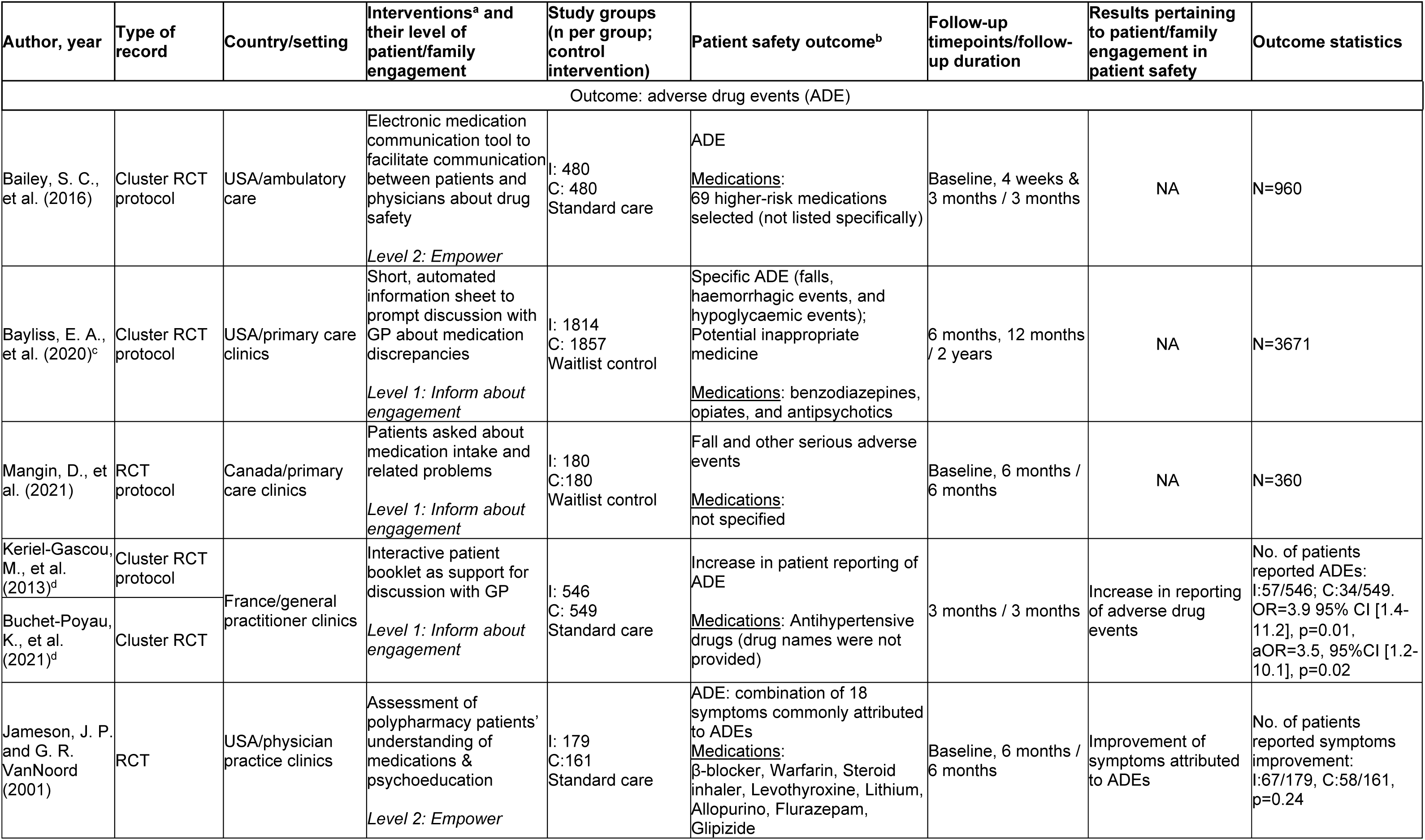
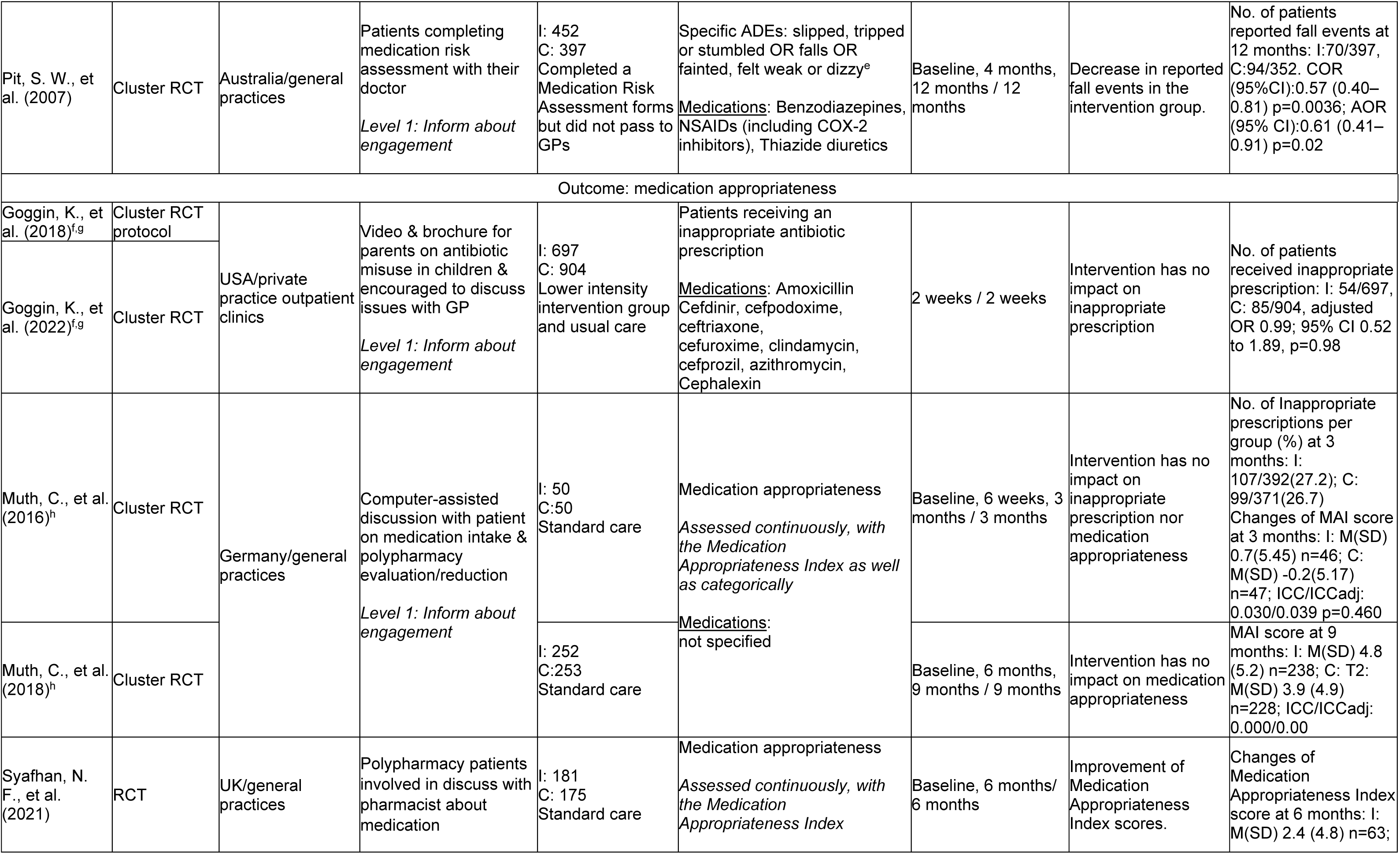
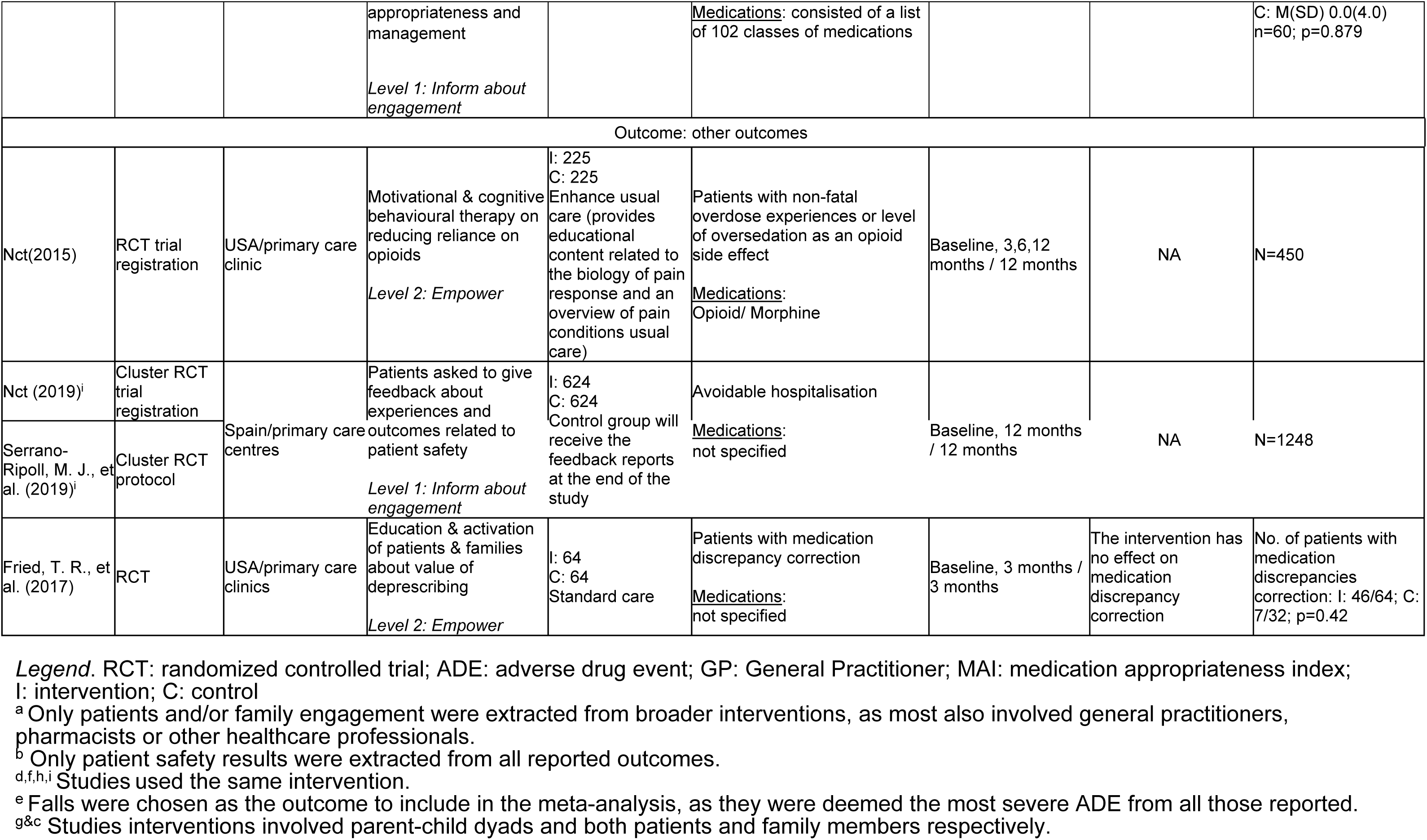
Study characteristics.

All included records were published between 2001 and 2021 (Table 2). Eight records presented completed studies, of which five were cluster Randomized Controlled Trials (RCTs) and three were standard RCTs. Of the remaining eight records, six were RCT protocols, and two were trial registration records.

Seven records from the United States, two from Germany, two from France, two from Spain (both reporting on the same project), one each from, respectively, Canada the United Kingdom, and Australia. The authors’ country of affiliation matched where projects were carried out for all completed RCTs.

The follow-up duration across RCTs ranged from two weeks to two years, and the number of randomized participants varied, with sample sizes ranging from 100 to 1,601 participants.

### 3.2 Outcome and intervention characteristics

Patient safety outcomes examined were predominantly adverse drug events (n = 8; three completed RCTs) and assessments of medication appropriateness (n = 8; six completed RCTs) (Table 2). One RCT protocol (36) listed both outcomes. Avoidable hospitalizations were reported by two records, of which no completed RCTs (37, 38).

The included 16 records described 12 types of interventions (Table 2). In terms of levels of patient and family engagement, most interventions remained at the *Inform about engagement* level (n=8), a few were at the *Empower* level (n=4), and none reached the *Partner or Integrate* level (Figure 2). There were proportionally more patient engagement interventions reaching Level 2 among study protocols and registrations (2/6, 33.3%) than among completed RCTs (2/8, 25%).

**Figure 2:**
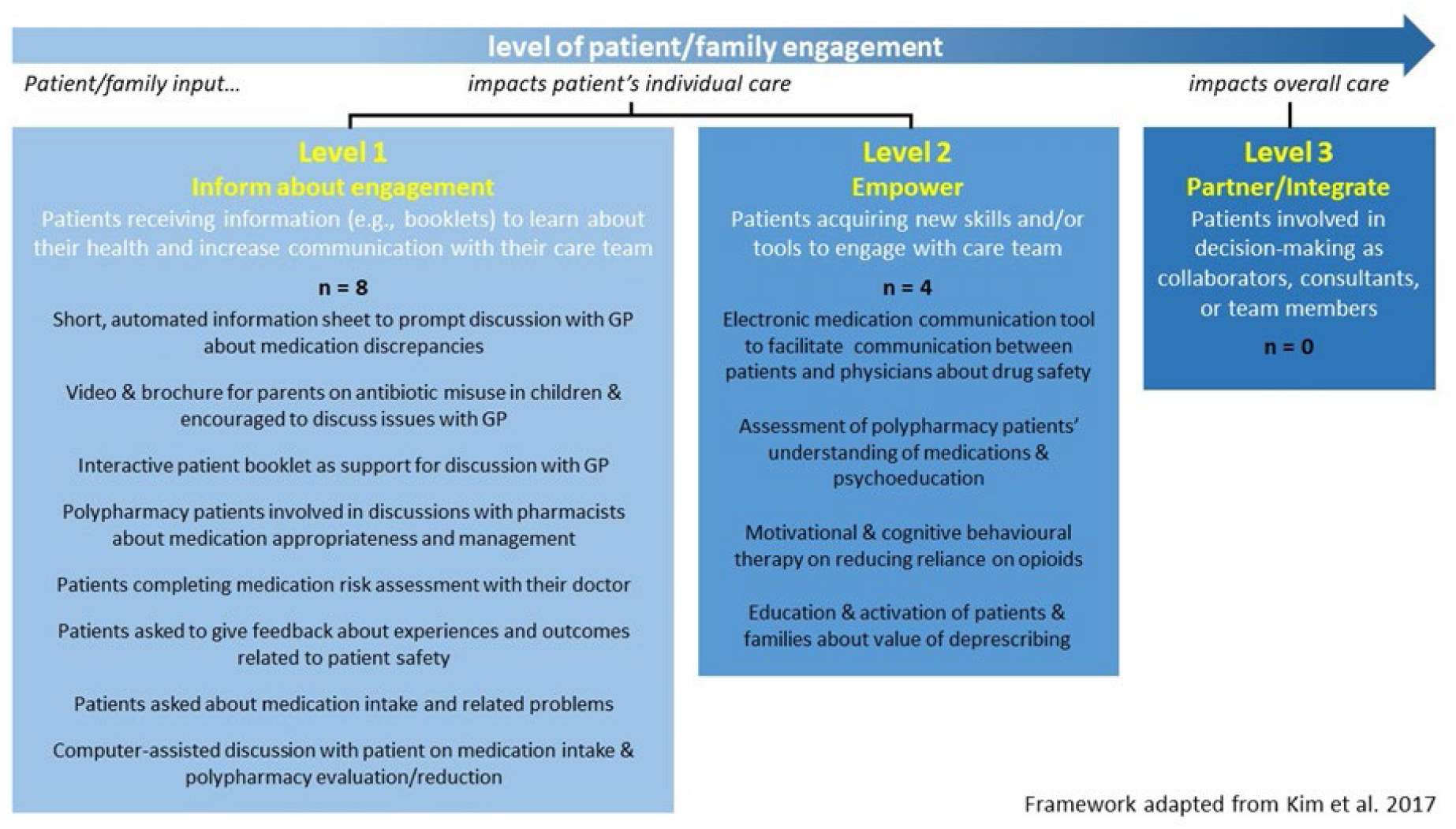
Interventions classified by level of patient and family engagement.

At the *Inform about engagement* level, most interventions (5/8, 62.5%) involved the provision of information to patients or their families to increase their understanding of health management. Patients partook in discussions with pharmacists about medication appropriateness and management (39), completed medication risk assessment forms were asked for feedback regarding safety outcomes during consultations (37,40), were asked about medication intake with or without the aid of a computer assistant (41, 42), or were provided information through video or brochures that served as basis for discussion with their GPs (36,43–45). At the *Empower* level, interventions included educational initiatives on safety deprescribing (45), online platforms fostering communication between patients and GPs regarding drug safety (46), motivational/cognitive-behavioural therapy aimed at reducing reliance on opioids (47), and psychoeducational support promoting polypharmacy patients’ understanding of their medication (48).

Two interventions described in three records included family members as participants: one involved education and activation strategies for patients and their families to promote deprescribing (36), while two records describing the same intervention reported the use of videos and brochures to promote communication between parents and general practitioners regarding antibiotic misuse in children (43,49).

### 3.3 Risk of bias assessment

Five RCTs demonstrated a relatively low overall risk of bias (Supplemental Figure 1). Two studies (40, 45) raised some concern, whereas one RCT (48) was assessed as having a high overall risk of bias.

### 3.4 Results of individual studies

Whereas most trial registrations and study protocols were either published within the previous two years or have already been followed by a publication on a corresponding RCT, none were found for one trial registration (47) and one published protocol (46) despite having been published eight to six years ago.

Of the eight completed RCTs, improvements were reported in all of those focusing on adverse drug events (all at Level 1 engagement; 40, 44, 48). Among completed RCTs focusing on medication appropriateness, only one out of four found a significant positive effect (Level 2 engagement; 38), whereas three other RCTs, all at Level 1 engagement, did not find significant effects (41,43,50). A single RCT, at Level 1 engagement, investigated medication discrepancy correction (47) and reported no significant changes following the intervention. Of note, however, intervention and outcome in this study were not fully aligned, as intervention consisted in patient and family activation and education about the value of deprescribing whereas the patient safety outcome was the correction of medication discrepancies.

### 3.5 Impact on specific outcomes

We conducted three separate meta-analyses to analyse the outcomes related to adverse drug events and medication appropriateness, which combined evidence from six out of eight completed RCTs (Figure 3). For medication appropriateness, the analysis was separated in categorical outcomes for studies reporting presence vs absence of inappropriate prescriptions and continuous outcomes for studies using the Medication Appropriateness Index. One study by Muth 2016 reported both measures and was therefore included in both analyses.

**Figure 3:**
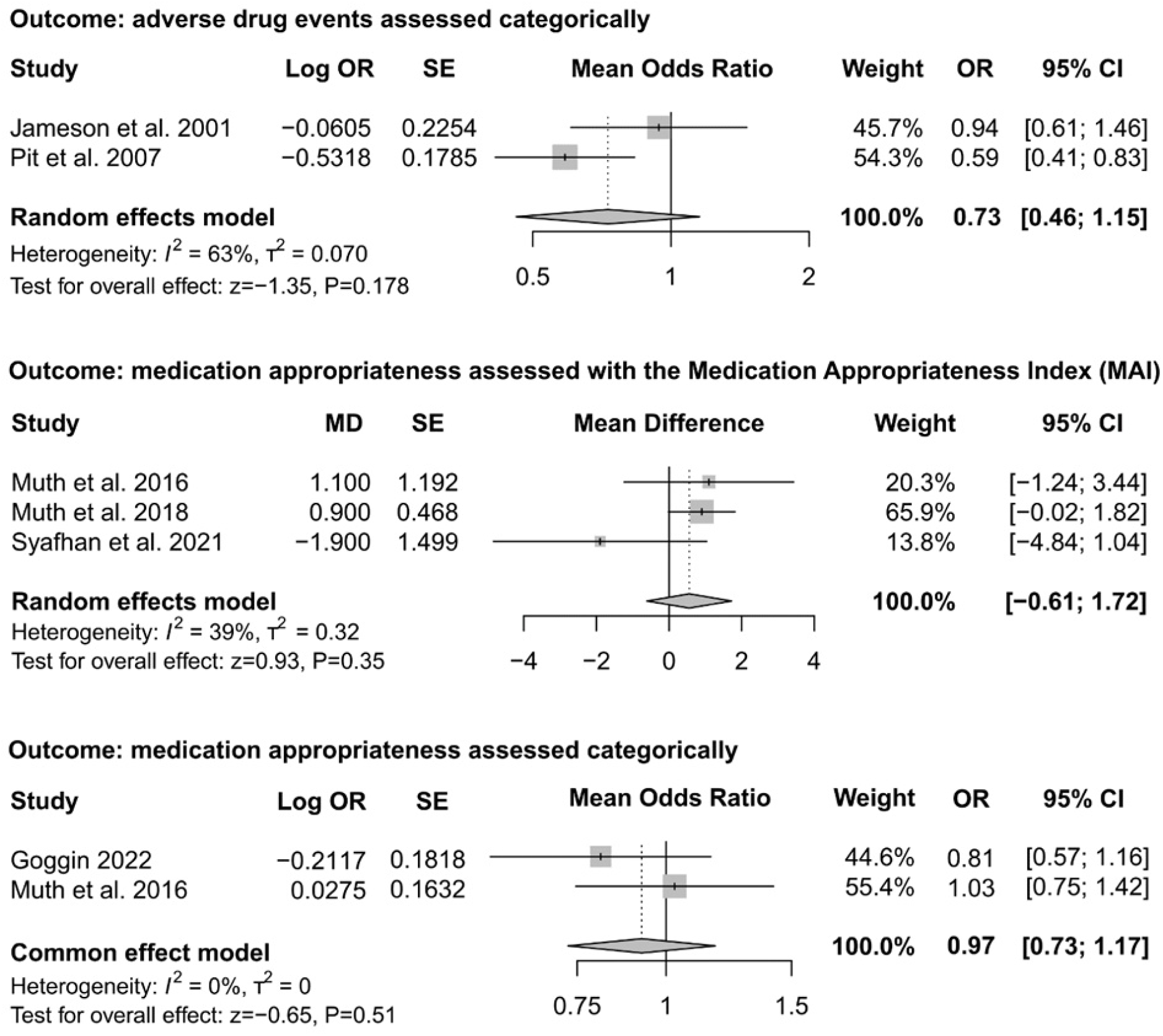
Meta-analyses of RCTs and cluster RCTs included in the review.

The meta-analysis on adverse drug events (Figure 3, upper panel) did not include one cluster RCT on the topic (46), as it was promoting self-reporting of adverse drug events (with more reported adverse drug events considered more favourable) whereas the two other studies assessed adverse drug events, with more events considered less favourable (40, 48). In the combined analysis, substantial heterogeneity was observed (I² = 63%) and the combined effect was non-significant (OR = 0.72, 95%CI [0.46, 1.15], p = 0.178). The certainty of the evidence for this meta-analysis was assessed as very low (Supplemental Table S6). Domains of concern during level of certainty assessment included study design, indirectness resulting from reported proxy events, and imprecision associated with wide confidence intervals.

The meta-analysis of the three studies reporting on medication appropriateness using the Medication Appropriateness Index Score (39, 41, 51; Figure 3, middle panel) found moderate heterogeneity (I² = 39%) and a non-significant mean difference of Medication Appropriateness Index score (MD = 0.56, 95%CI [-0.61, 1.72], p = 0.350). The certainty of evidence for this meta-analysis was rated as moderate (Supplemental Table 1). The downgrading of the level of evidence pertained to imprecision (broad confidence intervals).

The meta-analysis of categorically measured medication appropriateness (count of inappropriate prescriptions; Figure 3, lower panel) including two studies (43, 51) with low heterogeneity (I² = 0%) was also non-significant (OR = 0.97, 95%CI [0.73, 1.17], p = 0.514). The certainty of the evidence of this meta-analysis was high (Supplemental Table S6).

Medication discrepancy correction also had a moderate level of certainty based on the single study investigating this outcome (Supplemental Table S6).

Ultimately, none of the combined effect of interventions yielded significant results, although trends suggested beneficial effects for all outcomes considered.

## 4 Discussion

The present review identified 16 interventions aimed at promoting patient and family engagement in the context of patient safety within primary care settings. The scope of patient and family engagement remained limited, with none of these interventions offering patients and families the opportunity to influence level of overall care. All but one record focused on medication safety as an outcome. The meta-analyses conducted did not yield statistically significant combined effects, although approximately half of the completed RCTs reported modest to moderate positive effects of patient and family engagement interventions individually.

The lack of interventions at the global care level aligns with findings in broader healthcare settings finding no studies that have achieved the integration of patients as full care team members (34). The observed lack of effectiveness in certain interventions might also be attributed to inadequate statistical power, which could be associated with insufficient follow-up durations or small sample sizes. This limitation might have been particularly pronounced in studies investigating relatively infrequent patient safety outcomes, such as falls. Additionally, the intervention had a considerable overlap with standard of care. Many of them offered one-time consultations or written information, which, while potentially useful in identifying certain existing safety issues, may be insufficient or too short-term to provide more important shifts in the mindsets or behaviours of patients and their families.

The diversity of the interventions was limited, and except for medication reconciliation, the evidence-based strategies of patient and family engagement recommended by the Agency for Healthcare Research and Quality (52) were not investigated. Examples of such engagement strategies include being prepared to being engaged (patients and families encouraged to prepare for their appointments), teach-back (asking the patient/family to explain the instructions in their own words), and warm handoff (in-person handoff conducted in front of the patient).

Further, family involvement in the reported interventions remained limited to only three studies, two of which occurred in a paediatric setting (43, 49). Although the incorporation of family members introduces complexities in terms of study design, trials can be adapted to accommodate the needs of both patients and families, e.g., by providing separate study information materials or using modified surveys for family members. Meanwhile, family engagement remains a valuable resource in routine clinical practice, where research has demonstrated its potential to enhance communication between patients and providers, as evidenced by longer consultation times and patients taking a more active role during consultations (50).

Our review underscores the dearth of research into safety outcomes in primary care beyond the scope of medication safety. In particular, errors linked to other aspects of primary care delivery, such as communication errors or errors associated with care management may be important to target (7). Such errors occur at a high frequency (53), being estimated at 4 out of every 1000 primary care encounters (54).

The current review is the first to provide a comprehensive overview of randomised controlled interventions targeting patient and family engagement in primary care patient safety. It benefits from a comprehensive approach, including not only completed RCTs but also trial registrations and protocols, and a rigorous methodology, adhering to the Cochrane Collaboration guidelines at each step.

Nonetheless, this work has several limitations. Despite the search strategy being based on existing systematic reviews on patient safety and having been refined with a university librarian’s guidance, it may have omitted keywords relevant to patient safety outcomes different from medication safety. It has been pointed out by other authors that varying definitions of adverse events in primary care may lead to an underrepresentation of less commonly recognised or documented adverse events in the literature (55). Similarly, the absence of a universally recognised definition of patient engagement may have resulted in the potential exclusion of pertinent records due to terminological variations. Our search strategy, restricted to English-language, peer-reviewed publications on RCTs, may have omitted relevant records in other languages or on studies with less resource-expensive designs.

Overall, patient and family engagement is underutilized but shows promise. Investing efforts to bring such interventions to higher engagement levels and broader applications could make an impactful difference in primary care patient safety. There is also a compelling need to consider the inclusion of family members into the patient safety framework in all primary care settings, not only paediatrics.

## 5 Conclusions

Despite the potential of patient and family engagement in enhancing patient safety in primary care, there is a notable scarcity of studies, with the available evidence falling short of demonstrating unequivocal effectiveness. To extend interventions beyond health promotion and education about medication safety, future research may need to think outside of the box of traditional engagement approaches. Partnering up with patients and families during research design can be a relevant first step in this regard.

## 6 Declaration of Conflicting Interests

The author(s) declared no potential conflicts of interest with respect to the research, authorship, and/or publication of this article.

## Data Availability

All data produced in the present study are available upon reasonable request to the authors.

https://osf.io/sbacp

## 7 Funding

This study was funded by the Technology and Compassion: improving patient outcomes through data analytics and patientś voice in Primary Care. Funds from the National Medical Research Council (NMRC) Clinical Research Coordinator Funding were allocated to sustain the remuneration of both YP and FLL.

## 8 Role of the funder/sponsor

The funding entities played no part in shaping or executing the study, gathering, overseeing, analyzing, or interpreting the data, preparing, reviewing, or endorsing the manuscript, nor in the decision to submit the manuscript for publication.

## SUPPLEMENT FOR Patient and family engagement interventions in primary care patient safety: a systematic review and meta-analysis Complete search strategy

**Supplemental Table S1.**
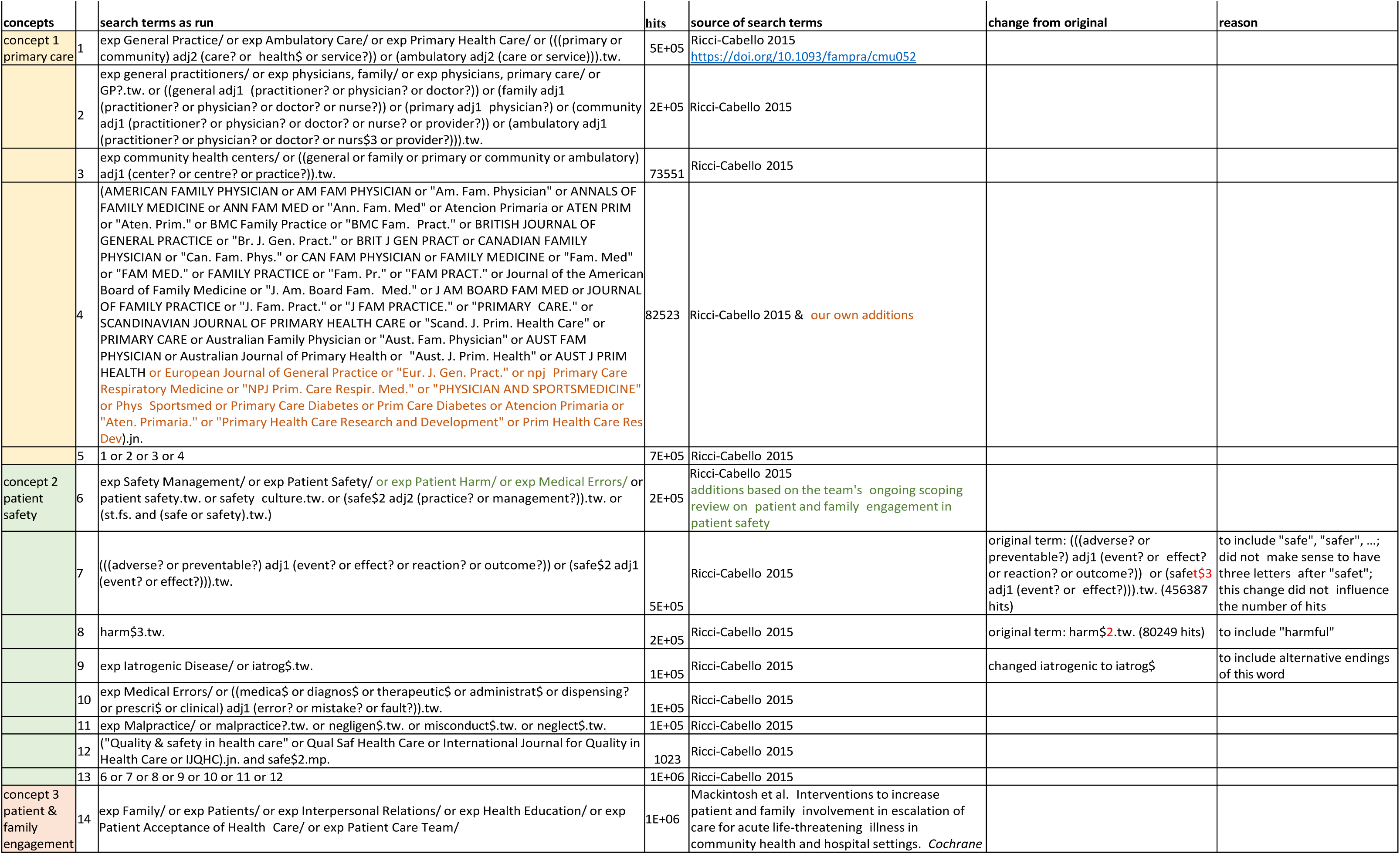
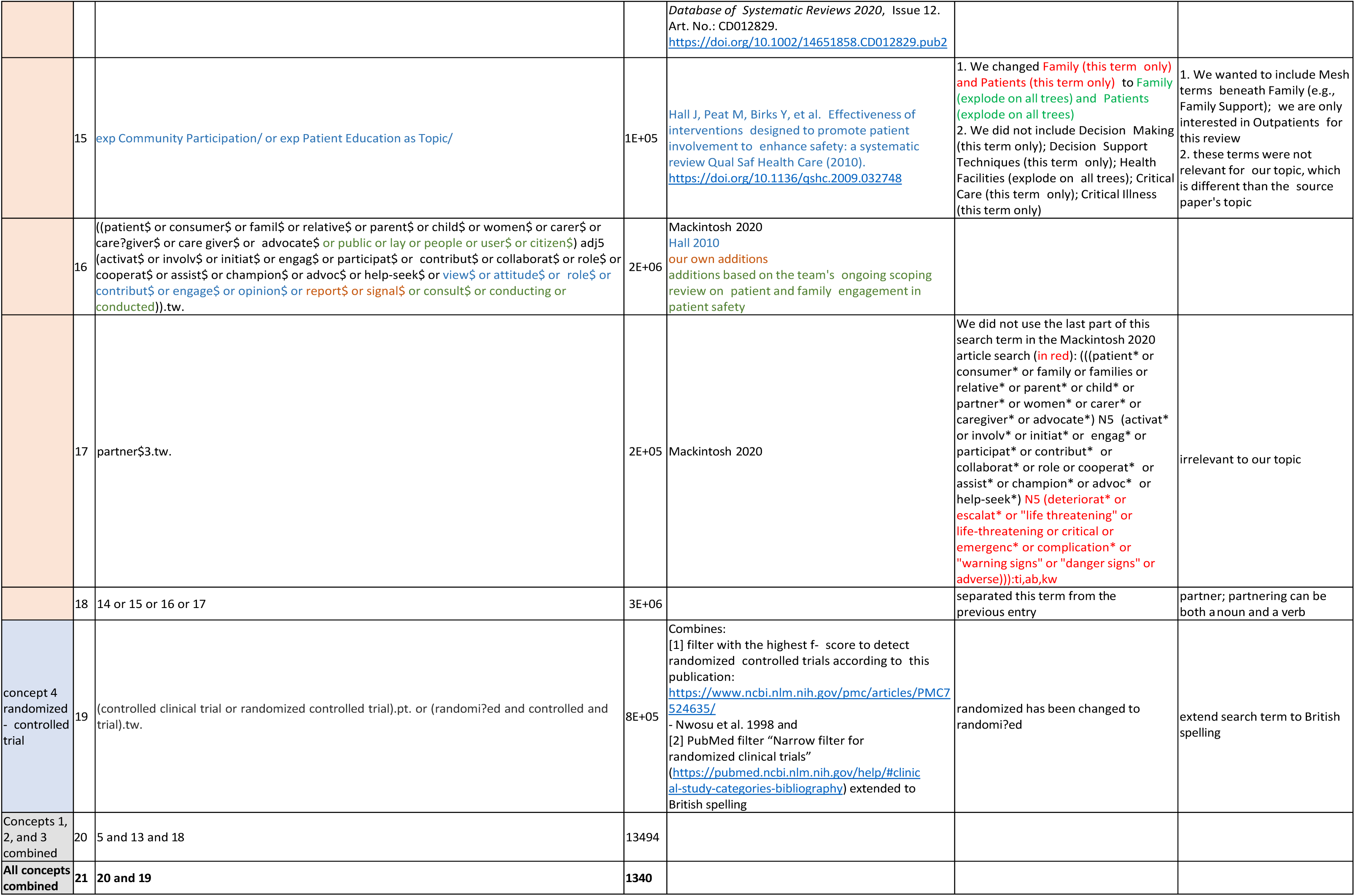
Ovid MEDLINE.

**Supplemental Table S2.** Embase.

| concepts |  | search terms as run |
| --- | --- | --- |
| concept 1<br>primary care | 1 | General Practice'/exp OR 'Ambulatory Care'/exp OR 'Primary Health Care'/exp OR (((primary OR community) NEAR/2 (care? OR health* OR service?)) OR (ambulatory NEAR/2 (care OR service))):ab,ti |
|  | 2 | general practitioners'/exp OR 'physicians, family'/exp OR 'physicians, primary care'/exp OR GP?:ab,ti OR ((general NEAR/1 (practitioner? OR physician? OR doctor?)) OR (family NEAR/1 (practitioner? OR physician? OR doctor? OR nurse?)) OR (primary NEAR/1 physician?) OR (community NEAR/1 (practitioner? OR physician? OR doctor? OR nurse? OR provider?)) OR (ambulatory NEAR/1 (practitioner? OR physician? OR doctor? OR nurse* OR provider?))):ab,ti |
|  | 3 | community health centers'/exp OR ((general OR family OR primary OR community OR ambulatory) NEAR/1 (center? OR centre? OR practice?)):ab,ti |
|  | 4 | ('AMERICAN FAMILY PHYSICIAN' OR 'AM FAM PHYSICIAN' OR 'Am. Fam. Physician' OR 'ANNALS OF FAMILY MEDICINE' OR 'ANN FAM MED' OR 'Ann. Fam. Med' OR 'Atencion Primaria' OR 'ATEN PRIM' OR 'Aten. Prim.' OR 'BMC Family Practice' OR 'BMC Fam. Pract.' OR 'BRITISH JOURNAL OF GENERAL PRACTICE' OR 'Br. J. Gen. Pract.' OR 'BRIT J GEN PRACT' OR 'CANADIAN FAMILY PHYSICIAN' OR 'Can. Fam. Phys.' OR 'CAN FAM PHYSICIAN' OR 'FAMILY MEDICINE' OR 'Fam. Med' OR 'FAM MED.' OR 'FAMILY PRACTICE' OR 'Fam. Pr.' OR 'FAM PRACT.' OR 'Journal of the American Board of Family Medicine' OR 'J. Am. Board Fam. Med.' OR 'J AM BOARD FAM MED' OR 'JOURNAL OF FAMILY PRACTICE' OR 'J. Fam. Pract.' OR 'J FAM PRACTICE.' OR 'PRIMARY CARE.' OR 'SCANDINAVIAN JOURNAL OF PRIMARY HEALTH CARE' OR 'Scand. J. Prim. Health Care' OR 'PRIMARY CARE' OR 'Australian Family Physician' OR 'Aust. Fam. Physician' OR 'AUST FAM PHYSICIAN' OR 'Australian Journal of Primary Health' OR 'Aust. J. Prim. Health' OR 'AUST J PRIM HEALTH' OR 'European Journal of General Practice' OR 'Eur. J. Gen. Pract.' OR 'npj Primary Care Respiratory Medicine' OR 'NPJ Prim. Care Respir. Med.' OR 'PHYSICIAN AND SPORTSMEDICINE' OR 'Phys Sportsmed' OR 'Primary Care Diabetes' OR 'Prim Care Diabetes' OR 'Aten. Primaria.' OR 'Primary Health Care Research AND Development' OR 'Prim Health Care Res Dev')/jt |
|  | 5 | 1 OR 2 OR 3 OR 4 |
| concept 2<br>patient safety | 6 | Safety Management'/exp OR 'Patient Safety'/exp OR 'Patient Harm'/exp OR 'Medical Errors'/exp OR 'patient safety':ab,ti OR 'safety culture':ab,ti OR (safe* NEAR/2 (practice? OR management?)):ab,ti |
|  | 7 | ((adverse? OR preventable?) NEAR/1 (event? OR effect? OR reaction? OR outcome?)) OR (safe* NEAR/1 (event? OR effect?)):ab,ti |
|  | 8 | harm*:ab,ti |
|  | 9 | Iatrogenic Disease'/exp OR iatrog*:ab,ti |
|  | 10 | Medical Errors'/exp OR ((medica* OR diagnos* OR therapeutic* OR administrat* OR dispensing? OR prescri* OR clinical) NEAR/1 (error? OR mistake? OR fault?)):ab,ti |
|  | 11 | Malpractice'/exp OR malpractice?:ab,ti OR negligen*:ab,ti OR misconduct*:ab,ti OR neglect*:ab,ti |
|  | 12 | ('Quality & safety in health care' OR 'Qual Saf Health Care' OR 'International Journal for Quality in Health Care' OR IJQHC)/jt AND safe*:ab,ti |
|  | 13 | 6 OR 7 OR 8 OR 9 OR 10 OR 11 OR 12 |
| concept 3<br>patient & family<br>engagement | 14 | family'/exp OR 'patients'/exp OR 'interpersonal relations'/exp OR 'health education'/exp OR 'patient acceptance of health care'/exp OR 'patient care team'/exp |
|  | 15 | Community Participation'/exp OR 'Patient Education as Topic'/exp |
|  | 16 | ((patient* OR consumer* OR famil* OR relative* OR parent* OR child* OR women* OR carer* OR care?giver* OR 'care giver' OR 'care givers' OR advocate* OR public OR lay OR people OR user* OR citizen*) NEAR/5 (activat* OR involv* OR initiat* OR engag* OR participat* OR collaborat* OR cooperat* OR assist* OR champion* OR advoc* OR help?seek* OR view* OR attitude* OR role* OR contribut* OR engage* OR opinion* OR report* OR signal* OR consult* OR conducting OR conducted)):ab,ti |
|  | 17 | partner*:ab,ti |
|  | 18 | 14 OR 15 OR 16 OR 17 |
| concept 4<br>randomized-<br>controlled trial | 19 | ('controlled clinical trial' OR 'randomized controlled trial'):pt OR (randomi?ed AND controlled AND trial):ab,ti |
| Concepts 1, 2,<br>and 3 combined | 20 | 5 AND 13 AND 18 |
| All concepts<br>combined | 21 | 20 AND 19 |

**Supplemental Table S3.**
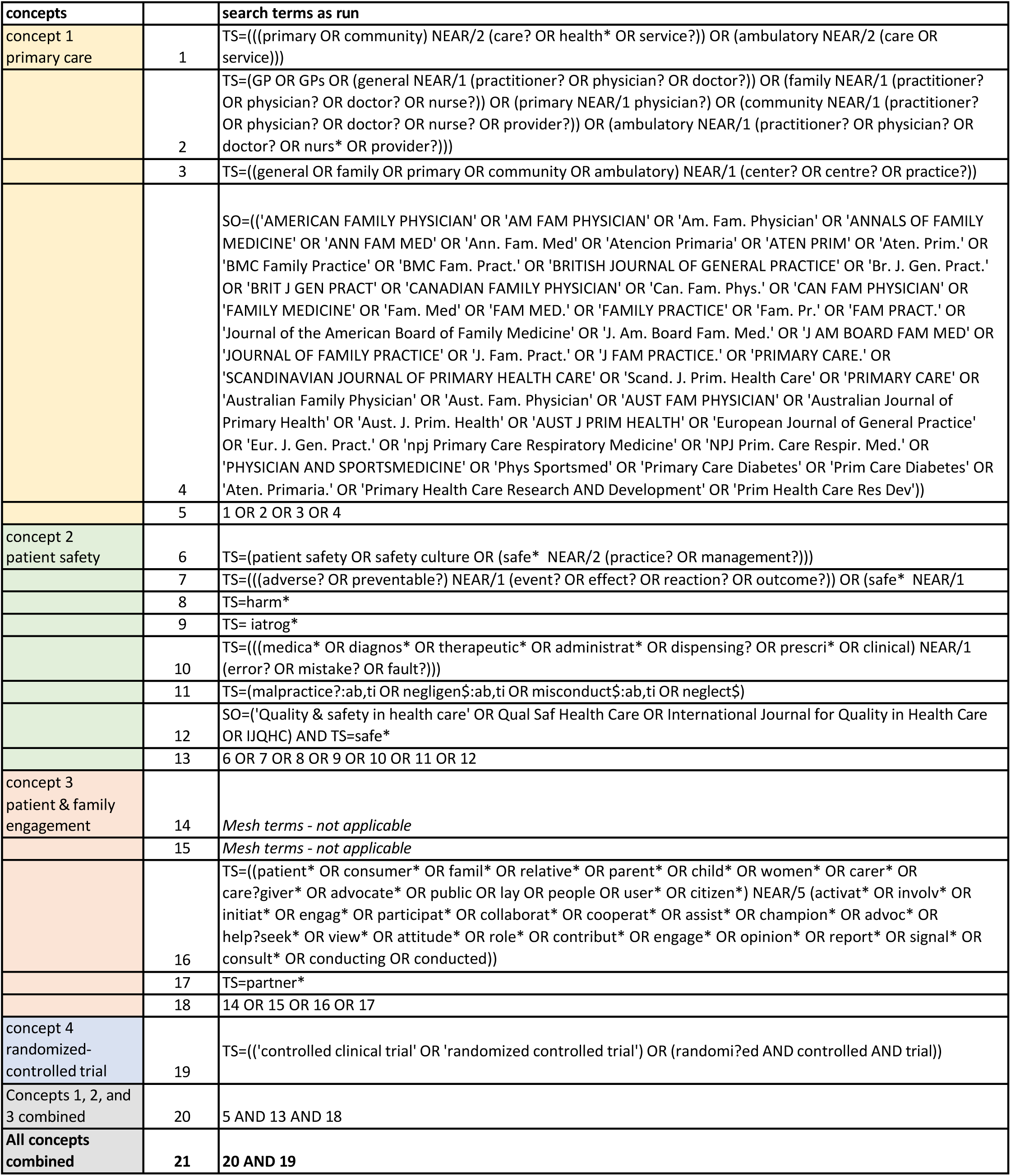
Web of Science.

**Supplemental Table S4.**
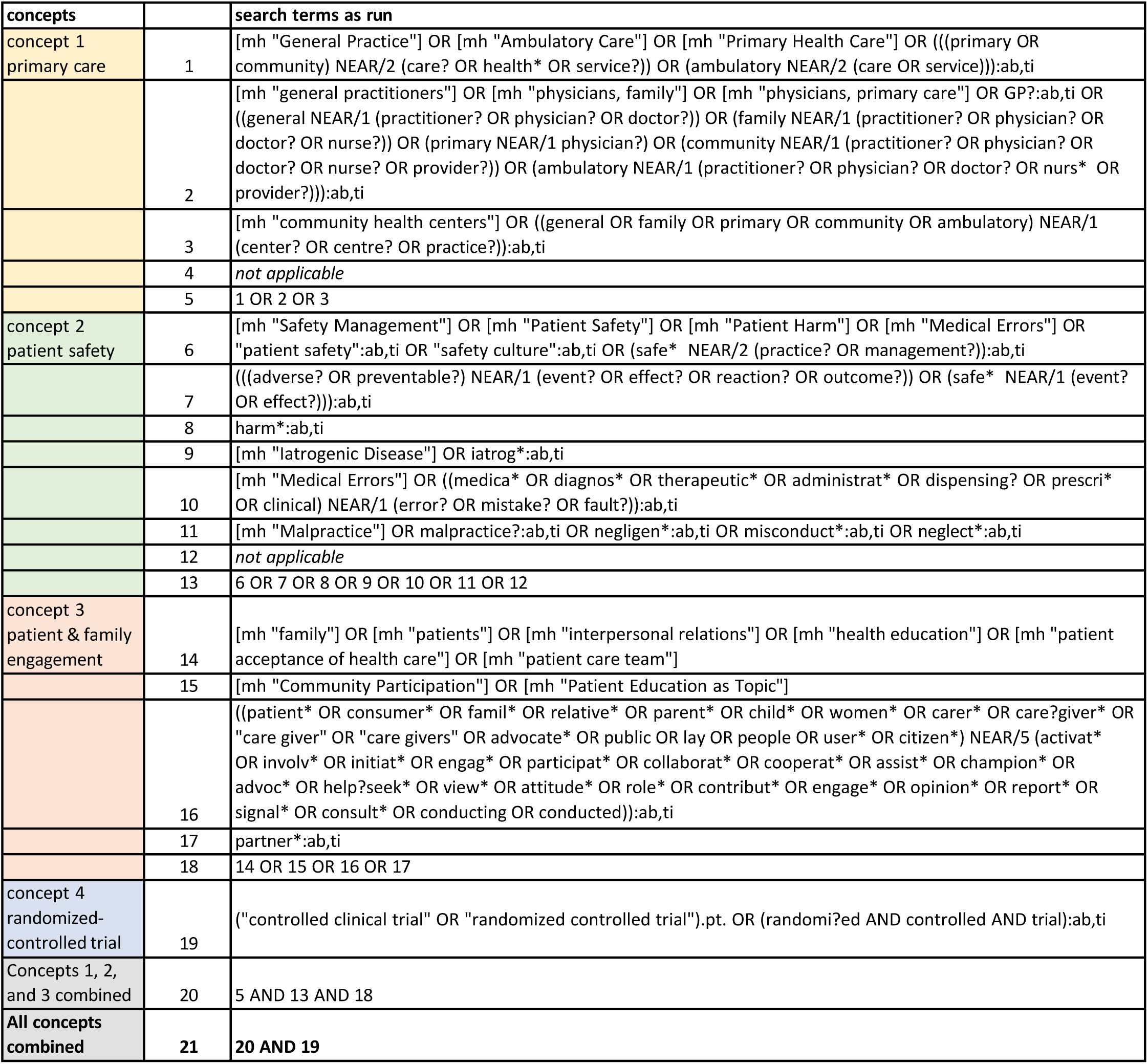
Cochrane.

**Supplemental Table S5.** CINHAL.

| concepts |  | search terms as run |
| --- | --- | --- |
| concept 1<br>primary care | 1 | MH "General Practice+" OR MH "Ambulatory Care+" OR MH "Primary Health Care+" OR (((primary OR community) N2 (care# OR health* OR service#)) OR (ambulatory N2 (care OR service))):ab,ti |
|  | 2 | MH "general practitioners+" OR MH "physicians, family+" OR MH "physicians, primary care+" OR GP#:ab,ti OR ((general N1 (practitioner# OR physician# OR doctor#)) OR (family N1 (practitioner# OR physician# OR doctor# OR nurse#)) OR (primary N1 physician#) OR (community N1 (practitioner# OR physician# OR doctor# OR nurse# OR provider#)) OR (ambulatory N1 (practitioner# OR physician# OR doctor# OR nurse* OR provider#))):ab,ti |
|  | 3 | MH "community health centers+" OR ((general OR family OR primary OR community OR ambulatory) N1 (center# OR centre# OR practice#)):ab,ti |
|  | 4 | *not applicable* |
|  | 5 | 1 OR 2 OR 3 |
| concept 2<br>patient safety | 6 | MH "Safety Management+" OR MH "Patient Safety+" OR MH "Patient Harm+" OR MH "Medical Errors+" OR "patient safety":ab,ti OR "safety culture":ab,ti OR (safe* N2 (practice? OR management#)):ab,ti |
|  | 7 | ((adverse# OR preventable#) N1 (event# OR effect# OR reaction# OR outcome#)) OR (safe* N1 (event# OR effect#)):ab,ti |
|  | 8 | harm*:ab,ti |
|  | 9 | MH "Iatrogenic Disease+" OR iatrog*:ab,ti |
|  | 10 | MH "Medical Errors+" OR ((medica* OR diagnos* OR therapeutic* OR administrat* OR dispensing# OR prescri* OR clinical) N1 (error# OR mistake# OR fault#)):ab,ti |
|  | 11 | MH "Malpractice+" OR malpractice#:ab,ti OR negligen*:ab,ti OR misconduct*:ab,ti OR neglect*:ab,ti |
|  | 12 | *not applicable* |
|  | 13 | 6 OR 7 OR 8 OR 9 OR 10 OR 11 OR 12 |
| concept 3<br>patient & family<br>engagement | 14 | MH "family+" OR MH "patients+" OR MH "interpersonal relations+" OR MH "health education+" OR MH "patient acceptance of health care+" OR MH "patient care team+" |
|  | 15 | MH "Community Participation+" OR MH "Patient Education as Topic+" |
|  | 16 | ((patient* OR consumer* OR famil* OR relative* OR parent* OR child* OR women* OR carer* OR care#giver* OR "care giver" OR "care givers" OR advocate* OR public OR lay OR people OR user* OR citizen*) N5 (activat* OR involv* OR initiat* OR engag* OR participat* OR collaborat* OR cooperat* OR assist* OR champion* OR advoc* OR help#seek* OR view* OR attitude* OR role* OR contribut* OR engage* OR opinion* OR report* OR signal* OR consult* OR conducting OR conducted)):ab,ti |
|  | 17 | partner*:ab,ti |
|  | 18 | 14 OR 15 OR 16 OR 17 |
| concept 4<br>randomized-<br>controlled trial | 19 | ("controlled clinical trial" OR "randomized controlled trial").pt. OR (randomi?ed AND controlled AND trial):ab,ti |
| Concepts 1, 2,<br>and 3 combined | 20 | 5 AND 13 AND 18 |
| **All concepts**<br>**combined** | **21** | **20 AND 19** |

**Supplemental Figure 1:**
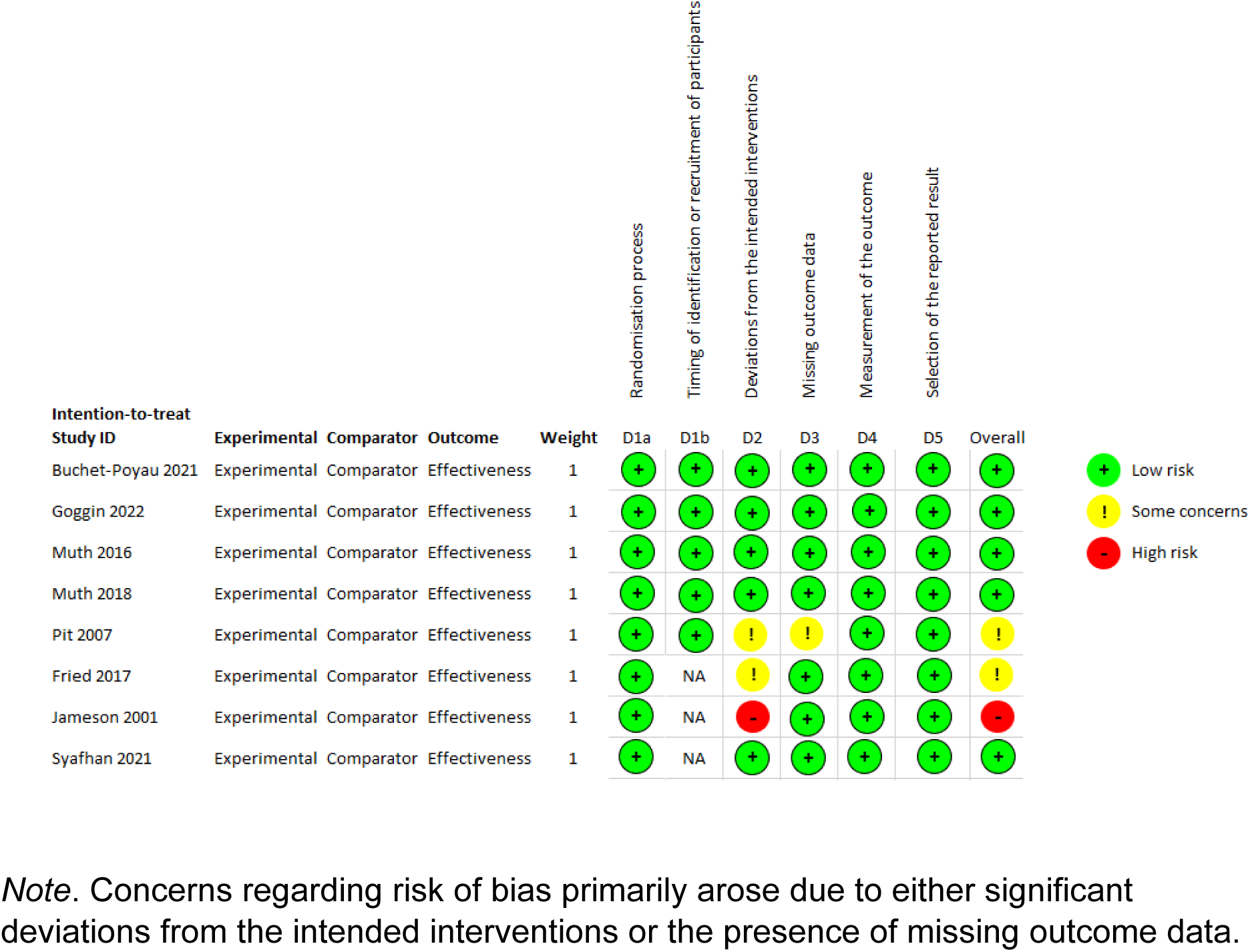
Risk of bias assessment of included cluster RCTs and RCTs.

**Supplemental Table S6:**
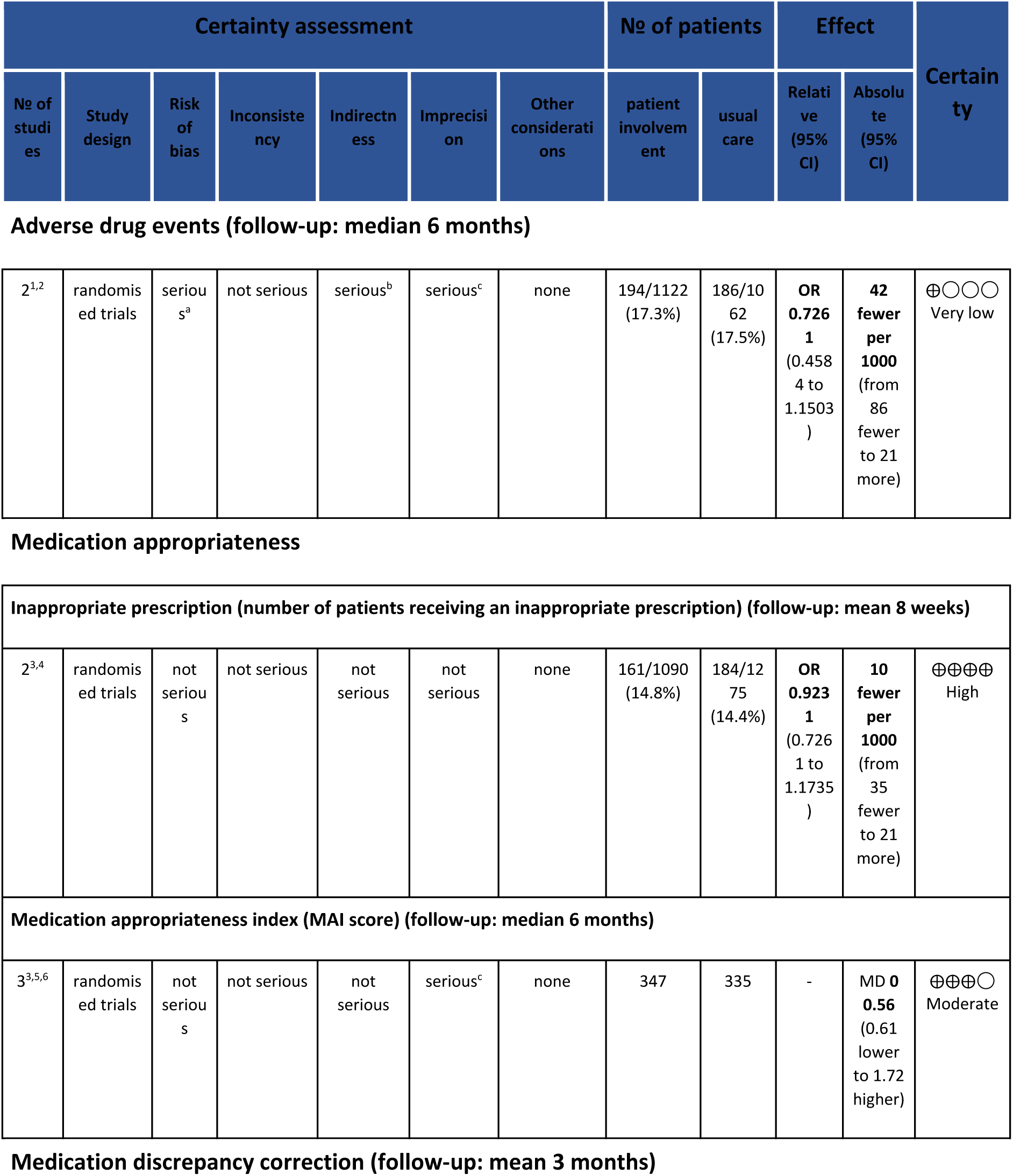
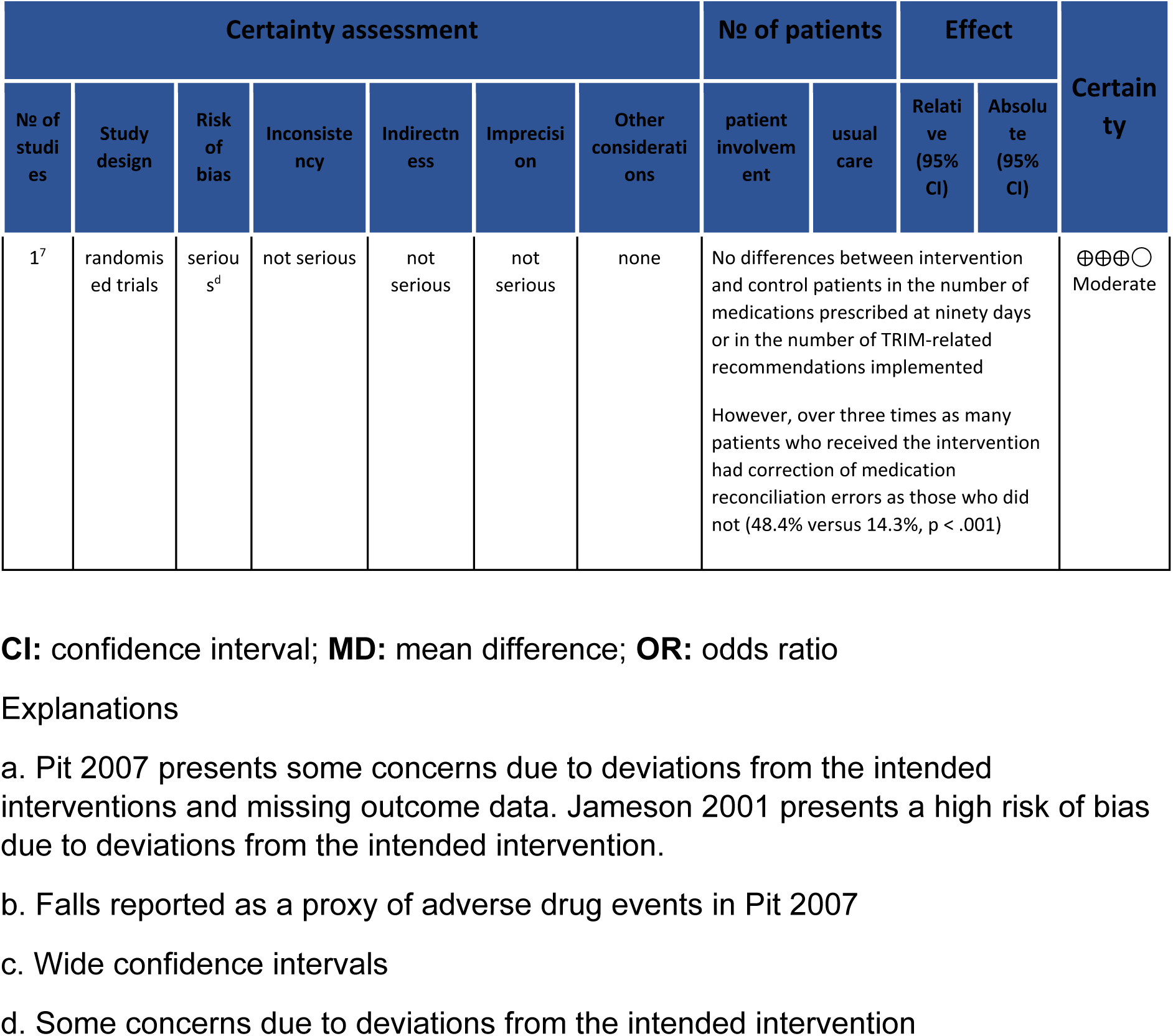
Patient and family engagement compared to standard of care to improve patient safety.

## Notes

### Competing Interest Statement

The authors have declared no competing interest.

### Clinical Protocols

https://www.crd.york.ac.uk/PROSPERO

### Funding Statement

This study was funded by Technology and Compassion improving patient outcomes through data analytics and patients voice in Primary Care

### Summary of Updates

We have shortened the words to about 3000 now. The meta-analysis results have been revised.

